# Atazanavir/ritonavir increased tizoxanide exposure from oral nitazoxanide through pharmacokinetic interaction in healthy volunteers

**DOI:** 10.1101/2023.09.20.23295544

**Authors:** Abdulafeez Akinloye, Timothy Oyedeji, Oluwasegun Eniayewu, Babatunde Adeagbo, Oluseye Bolaji, Steve Rannard, Andrew Owen, Adeniyi Olagunju

## Abstract

**Aims:** Nitazoxanide is a broad-spectrum antiviral with potential application in a number of viral infections. Its use is limited by gastrointestinal side effects associated with increasing dose. In this study, we investigated the possibility of enhancing the exposure of its active metabolite, tizoxanide, through pharmacokinetic interaction with atazanavir/ritonavir.

**Method:** This was a crossover drug-drug interaction study, 18 healthy participants received a single dose of 1000 mg of nitazoxanide alone in period 1 and in combination with 300/100 mg atazanavir/ritonavir in period 2 after a washout period of 21 days. On both days, blood samples for intensive pharmacokinetic analyses were collected before and at 0.25, 0.5, 1, 2, 4, 6, and 12 h after dose. To explore the utility of dried blood spots (DBS) as alternative to plasma for tizoxanide quantification, 50 µL of blood from some participants was spotted on DBS cards. Pharmacokinetic parameters were derived by non-compartmental analysis and compared between periods 1 and 2. The correlation between tizoxanide concentration in plasma and DBS was also evaluated.

**Results:** Co-administration of nitazoxanide with atazanavir/ritonavir resulted in a significant increase in tizoxanide plasma exposure. The geometric mean ratios (90% CI) of tizoxanide AUC_0-12h_, C_max_ and C_12h_ were 1.872 (1.870 – 1.875), 2.029 (1.99 – 2.07) and 3.14 (2.268 – 4.352) respectively, were all outside the 0.8 – 1.25 interval, implying clinically significant interaction. DBS concentration (%CV) was 46.3% (5.6%) lower than plasma concentrations, with a strong correlation (R = 0.89, P < 0.001). Similarly, DBS-derived plasma concentration and plasma concentrations displayed very strong correlation with linearity (R = 0.95, P<0.001).

**Conclusion:** Co-administration with atazanavir/ritonavir enhanced tizoxanide exposure with no report of adverse events in healthy volunteers.

## INTRODUCTION

Nitazoxanide, 2-(acetyloxy)-N-(5-nitro-2-thiazolyl) benzamide, is a thiazolide anti-infective agent initially developed for the treatment of veterinary helminthics [1, 2]. It has been studied for its anti-infective and more recently antineoplastic properties as well as activity on bone resorption [3-5]. It possesses a broad spectrum antimicrobial activity against a wide range of human pathogens, including human helminthics, anaerobic bacteria, and protozoans [6-8]. Nitazoxanide has been reported to have excellent activity against a few non-protozoan parasites such as the intestinal tapeworms *Hymenolepis nana* and *Trichuris trichiura* as well as against viruses, including enteric, human immunodeficiency virus, the Middle East respiratory syndrome, viral hepatitis, the rotaviruses and ebola virus [9-13]. Nitazoxanide is rapidly metabolism to its active metabolite tizoxanide which is highly bound to plasma proteins, and the inactive tizoxanide-glucuronide [14]. Recently, it was reported to possess *in-vitro* antiviral activity against SARS-CoV-2 and was investigated in several drug repurposing trials for COVID-19 management [15, 16].

However, efficacy against a new pathogen often requires systemic exposure levels that are not achievable at the approved dose [17]. In addition to the time lost to getting a new dose approved in a disease outbreak, tolerability may be compromised with higher doses. This has been reported for nitazoxanide whose clinical use is limited by severe gastrointestinal adverse events. These adverse events have been reported to be associated with increasing doses of nitazoxanide higher than 1000 mg that are capable of disrupting patients’ activities [18-20]. As a result, the recommended and currently approved dose is 500 mg twice daily but 1000 mg has been used in previous studies with higher tolerability [21].

Precedent exists for concomitant administration of a drug with another agent that inhibits or induces certain aspect(s) of its disposition pathway to improve systemic exposure, a strategy known as pharmacokinetic boosting. This has been applied to drugs used in the treatment of different disease such as HIV and cancer [22-26]. Interaction between nitazoxanide and atazanavir/ritonavir has not been reported. However, since viral protease inhibitors inhibit uridine 5’-diphospho-glucuronosyltransferase (UDP-glucuronosyltransferase, UGT) enzymes [27], atazanavir/ritonavir is expected to inhibit the glucuronidation of tizoxanide, potentially increasing its exposure. We conducted a single-dose study in healthy volunteers to explore this concept.

## METHODS

### Study Design and Participants

This was a single centre, open-label, two-period crossover study, with a 3-week washout period in between. In period 1, participants received a single dose of 1000 mg of nitazoxanide alone (two 500 mg tablets) in the morning after a regular local meal. Following a 3-week washout period, participants received a single dose of 1000 mg of nitazoxanide (two 500 mg tablets) along with a single dose of 300 mg/100 mg atazanavir/ritonavir (co-formulated tablet) in period 2. That is nitazoxanide plus atazanavir/ritonavir in the morning. The study protocol was reviewed by the Institute of Public Health, Obafemi Awolowo University Ile-Ife, Nigeria and approved on 16 December 2020 (IPH/OAU/12/1574). The study is registered on ClinicalTrials.gov (NCT05680792). Participants were recruited from the community of Obafemi Awolowo University Ile-Ife, and included healthy individuals who were at least 18 years old, able to understand the study information, non-smokers, and non-alcoholics. Individuals who were pregnant, breastfeeding, have taken any medication or coffee within 2 weeks of their participation, or allergic to nitazoxanide, atazanavir, or ritonavir were excluded. Only participants who agreed to participate after understanding the study procedure and who signed the informed consent form were enrolment.

### Sample Collection

Intensive pharmacokinetic sampling was conducted after an observed single dose in each of the two periods, consisting of collection of 2 mL venous blood into EDTA tubes at 0, 0.25, 0.5, 1, 2, 4, 6, and 12 h after dose. Plasma was separated by immediate centrifugation at 3000 rpm for 5 min at 4 C. Plasma samples were transferred into cryovials and stored at -70 C until analysis. To evaluate the potential utility of the dried blood spot (DBS) method and for its cross-validation against the plasma method, intensive pharmacokinetic DBS samples were collected from six individuals in period 1 and six individuals in period 2. Briefly, before plasma separation from venous blood from each time point as described above, 50 µL per spot was spotted within the marked circles of Whatman 903® Protein Saver card (GE Healthcare Life Sciences, New York, USA. The cards were allowed to dry at room temperature for 2 h before being packed into zip-lock bags containing desiccants and stored at -70 C until analysis. Both plasma and DBS samples were analysed within one week of collection.

### Bioanalysis of Study Samples

Quantification of tizoxanide in plasma and DBS was carried out using an earlier reported LC-MS/MS method [28], modified and partially validated as follows. The LC system consisted of Accela Open Autosampler and an Accela LC-Pump (Thermo Fischer Scientific, Hemel Hempstead, UK). Chromatographic separation was carried out on a reverse phase Fortis™ C_18_ column (3 µm particle size, 100 mm x 2.1 mm; Fortis Technologies Ltd, Neston, Cheshire, UK) with a 2 μm C_18_ Quest guard-column (Thermo Electron Corporation, Hemel Hempstead, Hertfordshire, UK). Gradient elution consisted of 10 mM ammonium formate in acetonitrile (mobile phase A) and 10 mM ammonium formate in water (mobile phase B) at a flow rate of 400 µL/min. The gradient program started with 5% of mobile phase A, maintained for 1 minute, and increased to 80% at 1.5 min. It was further increased to 95% at 4 min and returned to 5% at 6 min. The total run time was 6 min. Detection was on the TSQ Vantage (Thermo Electron Corporation, Hemel Hempstead, Hertfordshire, UK) with a heated electrospray 3ptimizati source operated in the negative 3ptimizati mode and selective reaction monitoring. Xcalibur™ was used for compound tuning and 3ptimization while the Lcquan™ (version 2.7.0, Thermo Fisher Scientific, Hemel Hempstead, UK) was used for sequence acquisition and data processing.

Calibration standards for tizoxanide plasma and DBS assay consisted of zero blank and 9 calibrator levels ranging from 50 to 20000 ng/mL, prepared by serial dilution of plasma working stock of tizoxanide with drug-free plasma from a healthy volunteer. Quality control (QC) samples included a lower limit of quantitation (LLOQ; 50 ng/mL), low-QC (LQC; 120 ng/mL), medium-QC (MQC; 8600 ng/mL) and high-QC (HQC; 18000 ng/mL), similarly prepared from a different working stock. All working stock solutions and plasma working stock were prepared from a 1 mg/mL stock solution prepared from tizoxanide reference compound (Toronto Research Chemicals Inc., Toronto, ON, Canada). Efavirenz (Selleck Chemicals LLC, Houston, USA) was used as internal standard.

Extraction of tizoxanide from the validation samples and study samples was carried out by the protein precipitation method using acetonitrile. Briefly, 100 µL of plasma was transferred into clean 7 mL extraction tubes, followed by the addition of 20 µL of internal standard (efavirenz; 750 ng/mL) and 20 µL of formic acid. 1000 µL of acetonitrile was then added and vortexed for 10 s to precipitate proteins. The extract was left to stand for 10 min and vortexed again for 10 s before centrifugation at 3500 rpm for 10 min at 4 C. 400 µL of the supernatant was transferred into 400 µL glass autosampler vials and the injection volume was 25 µL. For DBS, three 6 mm punches were taken from a spot per sample and transferred into 7 mL extraction tubes with screw caps. 500 µL of acetonitrile and 20 µL of internal standard (efavirenz; 750 ng/mL) were added and vortexed for 10 s. The tubes were left to stand for 30 min, and vortexed for 10 s every 10 min. The extracts were centrifuged at 4000 rpm for 15 min at 4 C and 400 µL of the supernatants were transferred into another clean 7 mL extraction tube. 20 µL of formic acid was added to the supernatant and the samples were vortexed again. 400 µL of the final extract was transferred into 400 µL glass autosampler vials and 25 µL was injected. At least 3 validation assay batches for precision and accuracy were run for each sample type, and additional assay batches for selectivity, recovery, matrix effect, and extraction efficiency.

Plasma and DBS samples obtained from the study participants were analysed in the same way as described above.

### Data Analysis

Bioanalytical method validation data were processed using Excel® (Microsoft Inc., Washington, USA) and assessed using FDA guidance [29], including %CV within ±15% for QCs (±20% for LLOQ) in the case of accuracy and precision assays. Tizoxanide pharmacokinetic parameters (AUC_0-12h_, C_max_, C_12_ and t_max_) from the concentration-time data for both periods were obtained by non-compartmental analysis using GraphPad Prism® (GraphPad Software, La Jolla, CA). In line with the FDA guidance, geometric mean ratios and their 90% Cis were calculated for AUC_0-12_, C_max_, and C_12_ to assess the clinical significance of any observed interaction [30]. For the cross validation of the plasma and DBS methods, linear regression was first used to describe the relationship between measured plasma and DBS concentrations. DBS-derived plasma concentrations were calculated from the equation DBS-derived plasma = [DBS_conc_/(1 – hct)]x0.999 [31], where DBS_conc_ is the concentration measured in DBS, hct is average hematocrit [32] and 0.999 [14, 33] is the plasma bound fraction of tizoxanide. Afterwards, DBS-derived plasma concentrations were compared with the paired plasma concentration using linear regression analysis.

## RESULTS

### Participant’s disposition and demographics

A total of 25 participants consented to participate in the study but 4 withdrew from the study due to illnesses, non-related to the study, such as malaria while 3 were withdrawn due to non-compliance with the study protocol. Eventually, 18 participants (males, n = 10 and females, n = 8) completed the study. The mean (±SD) age and mean weight (±SD) were 27.4 (6.9) years and 60.4 (12.8) kg, respectively. In both periods of the study, no participant reported any serious symptom or adverse effect.

### Method Validation

#### Linearity, accuracy, and precision of plasma and DBS assay

Linearity of the assay method for both plasma and DBS was established in more than 8 assays over the range of 50 – 20000 ng/mL, with all the calibration standards having a percentage bias falling below ±15%. A typical regression coefficient (r) was 0.9971. For plasma assay, accuracy and precision were 84.4 – 112.2% and 1.0 – 0.6%; while for DBS, they were 91.8 – 108.4% and 0.49 – 13.1% (Supplementary S1). All values were within the acceptance criteria from FDA guideline.

#### Recovery, matrix effect, and extraction proficiency

The mean (%CV) recoveries of tizoxanide from plasma at LLOQ, LQC, MQC, and HQC were 88.8% (4.6), 87.1% (3.8), 110.1% (7.6) and 100.9 (2.3), respectively, and the overall mean (%CV) recovery was 96.7% (11.2). The plasma matrix effect was between 103.1% and 112.7% with a %CV less than ±15. Also, mean extraction proficiency was 104.4 with %CV less than 15% for all QC levels.

#### Pharmacokinetics of tizoxanide and the effects of co-administration of atazanavir/ritonavir

In period 1 (nitazoxanide alone) and period 2 (nitazoxanide plus atazanavir/ritonavir), tizoxanide was undetectable in the plasma until after 30 min following drug administration, and the median (range) t_max_ was consistently at 4 (2 – 6) hrs (Figure 1). Geometric mean tizoxanide AUC_0-12h_ was 124967.7 ng.h/mL in period 1 compared with 233984.1 ng.h/mL in period 2, representing GMR of 1.872 (Table 1). C_max_ was 4375.7 ng/mL for period 1 compared with 8882.1 ng/mL for period 2 (GMR: 2.029), while C_12_ was 553.8 ng/mL period 1 compared with 1740.7 ng/mL for period 2 (GMR: 3.143). The values obtained for the parameters were outside the 0.80-1.25 range indicating clinical significance. Similarly, PK parameters obtained from DBS-derived plasma concentrations showed increased exposure from period 1 to period 2, but the folds increase were more than what were observed in plasma (Supplementary S2 and S3).

**Figure 1:**
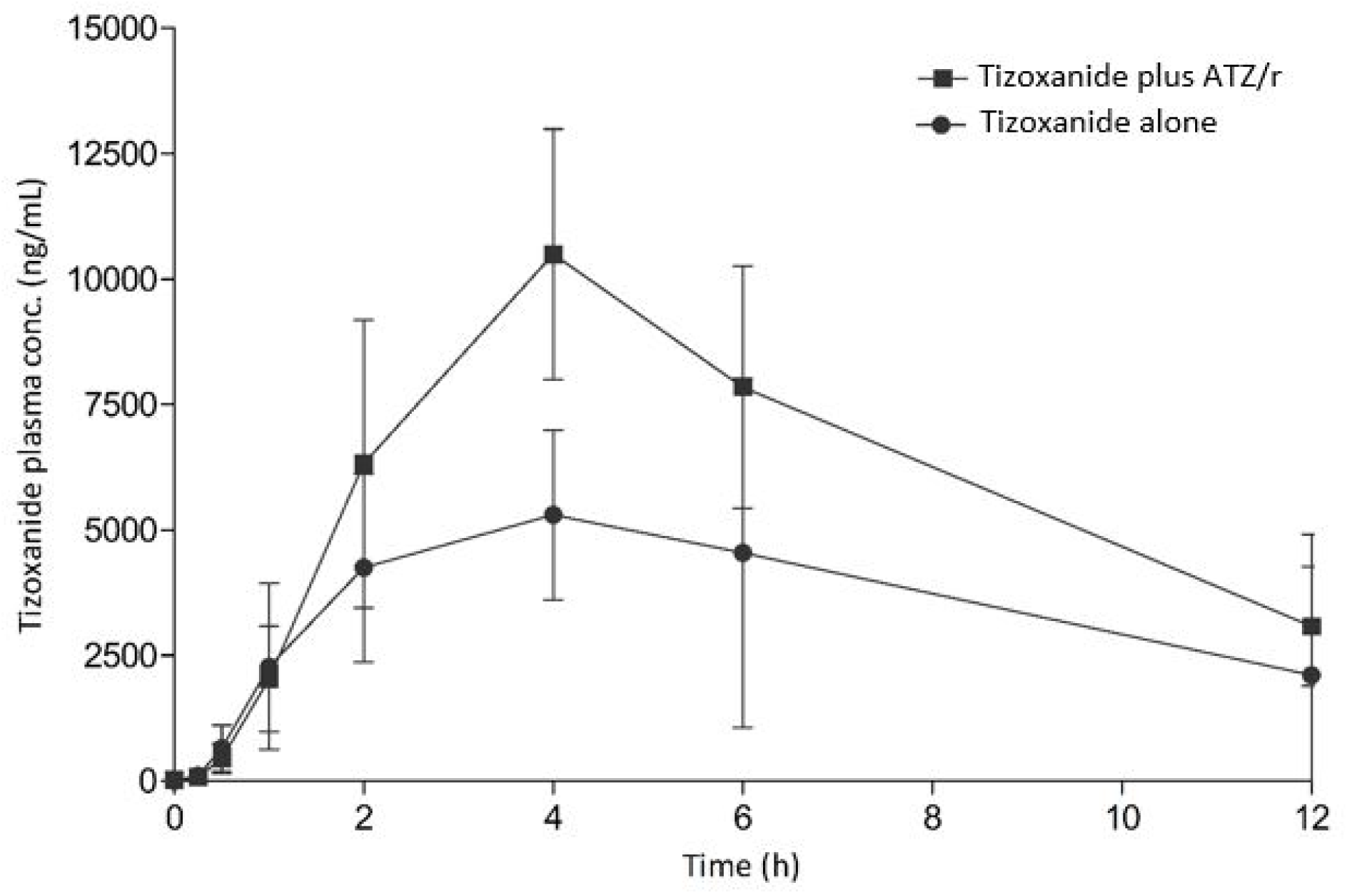
Plasma concentration-time curve of tizoxanide alone and with atazanavir/ritonavir (ATZ/r); data is presented here as mean (95% CI) for n =18.

**Table 1:**
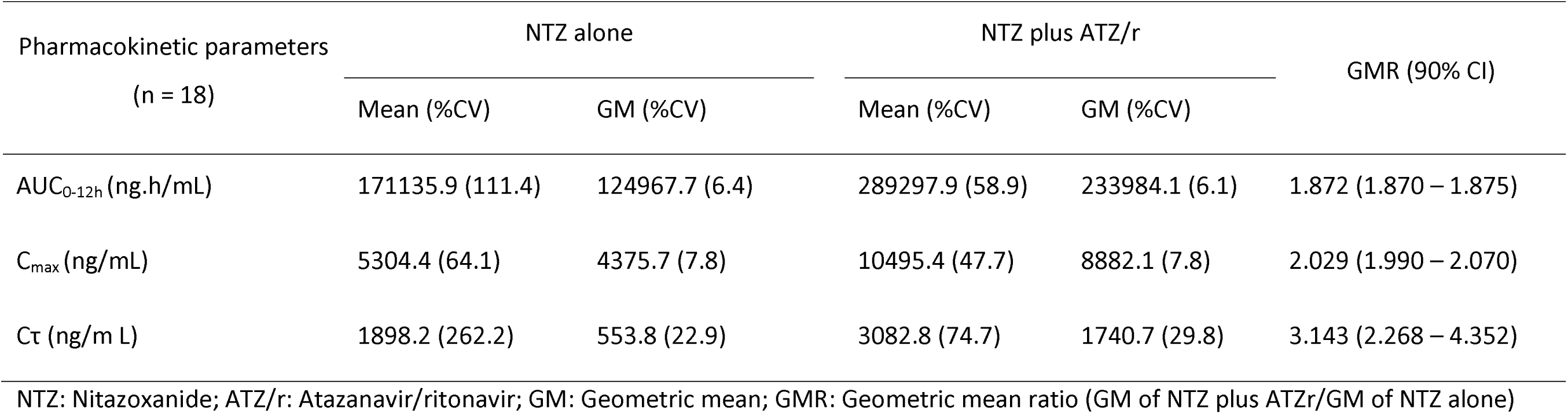
Pharmacokinetic parameters of plasma tizoxanide with and without atazanavir/ritonavir.

#### Plasma-DBS Cross-validation

Significant correlation was observed between nitazoxanide concentration in plasma and DBS correlation (coefficient, R = 0.87, P < 0.001) (Figure 2). The equation y = 2.1343x + 53.489 described the relationship between tizoxanide concentration in the two matrices, where y is the plasma concentrations and x is the DBS concentrations. Similarly, DBS-derived plasma concentrations obtained from DBS concentrations had a very strong correlation with measured plasma concentration with a correlation coefficient R = 0.95, P < 0.001. (Figure 3).

**Figure 2:**
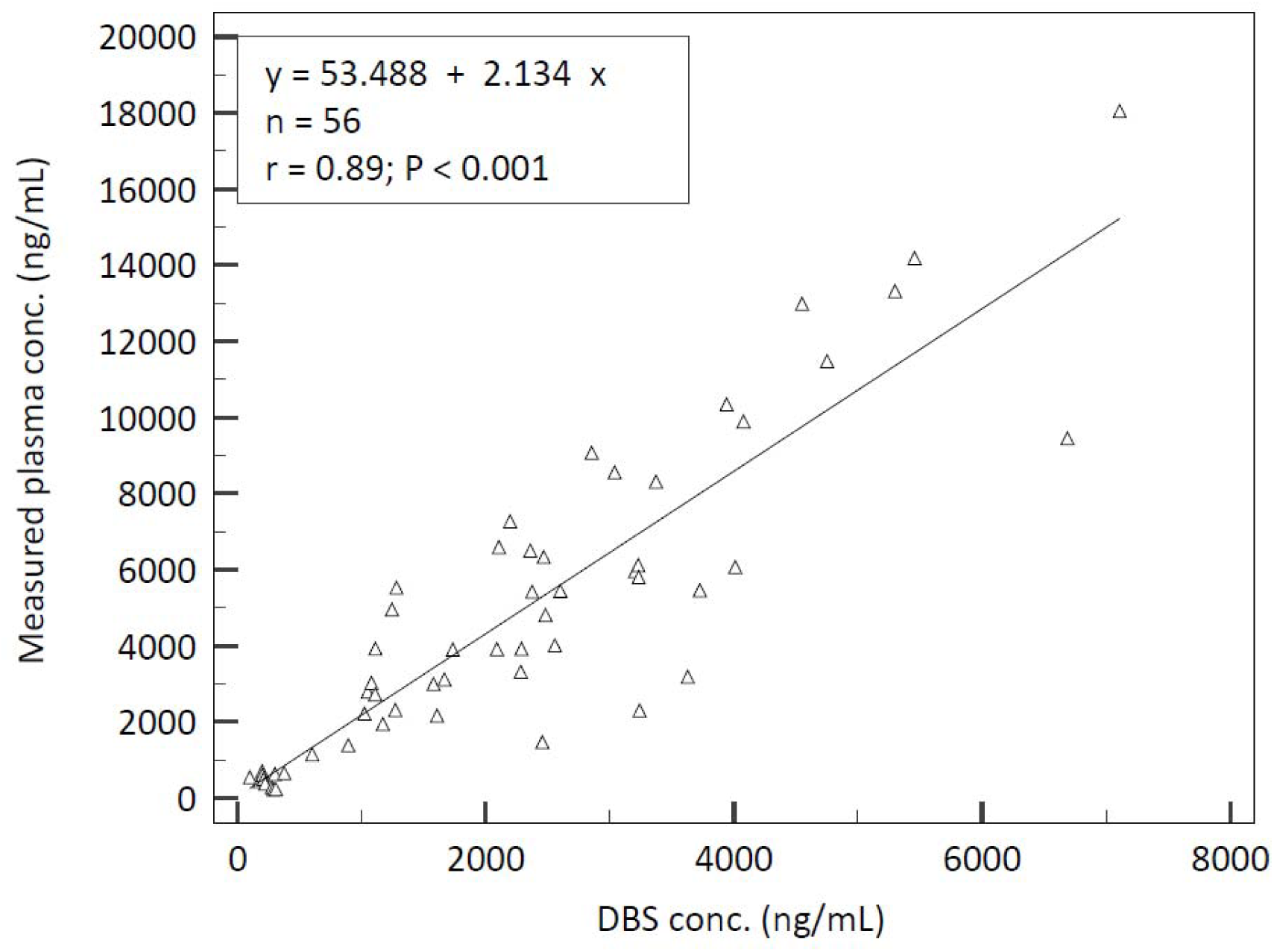
Linearity between tizoxanide plasma concentrations DBS concentrations.

**Figure 3:**
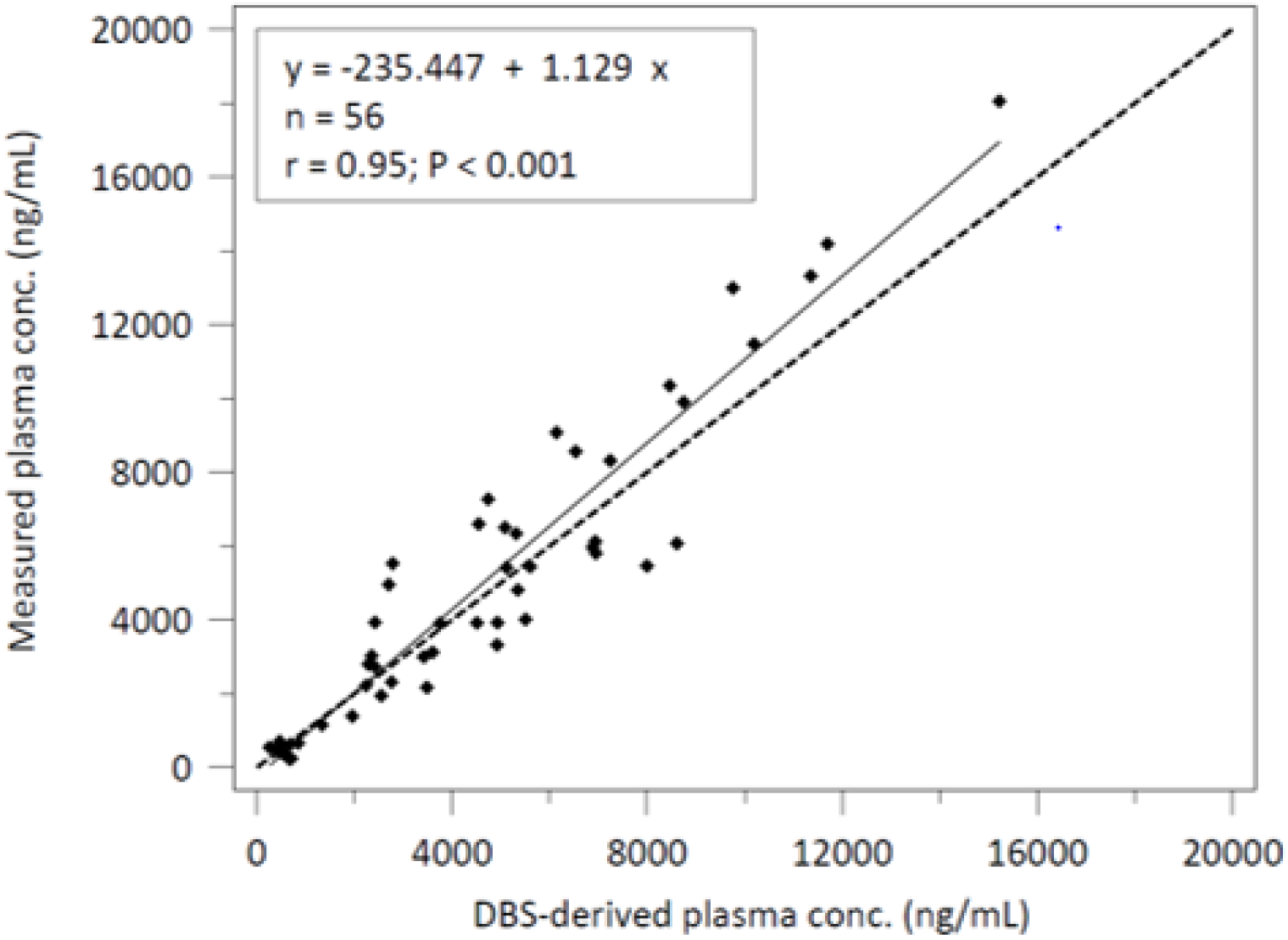
Linearity between tizoxanide plasma concentrations DBS concentrations (broken line represents true line of identity)

## DISCUSSIONS

The pharmacokinetic interaction between oral nitazoxanide (1000 mg) and atazanavir/ritonavir (300/100 mg) was successfully investigated in this single dose, cross-over study in healthy volunteers. Co-administration with atazanavir/ritonavir boosted the plasma pharmacokinetics of tizoxanide, the active metabolite of nitazoxanide, by 69.05%. This strategy could potentially create opportunities to explore nitazoxanide for indications requiring exposure levels not achievable with the standard dose, with further benefit of synergistic antiviral activity of atazanavir/ritonavir in the case of viral infections.

Doses higher than 500 mg of nitazoxanide were predicted to achieve tizoxanide plasma and lung tissue levels to ensure EC_90_ of 1.43 mg/L required for SARS-CoV-2 viral suppression [17]. Since efficacy, ease of administration and more importantly safety concerns usually inform the choice of drugs and dosing in repurposed drugs [34, 35], we decided to investigate the appropriateness of a 1000 mg twice daily regime since its safety has been previously established and not associated with most gastrointestinal side effects associated with higher doses [36, 37]. Although this dose was reported to only achieve plasma concentration higher than the EC_90_ and not in the lungs, the strategy investigated here resulted in the *in situ* elevation of tizoxanide metabolite, preventing its conversion to tizoxanide-glucuronide by UGT enzymes, in the presence of atazanavir, a known UGT enzyme inhibitor [27]. Additionally, atazanavir/ritonavir is an antiviral drug with activity against several viral pathogens; it is reportedly active *in vitro* against SARS-CoV-2 [19, 38, 39]. Hence, its combination with nitazoxanide could be investigated for efficacy against SARS-CoV-2 infection and other viruses in the future. Furthermore, this combination was not associated with any serious adverse events in this cohort.

Though atazanavir is a known inhibitor of UGT enzymes, investigating the exact mechanism was beyond the scope of this study. However, pharmacokinetic boosting by intentional modulation of drug disposition is not new and it is extensively used in HIV treatment (e.g. lopinavir/ritonavir, atazanavir/cobisistat, elvitegravir/ cobisistat). A recent review is also available on pharmacokinetic boosting strategies for kinase inhibitors [24].

Our exploration of the DBS method as an alternative for the plasma method was prompted by sample collection limitations associated with risk mitigation strategies in pandemic settings. Samples collected on DBS cards are considered non-infectious and are exempted from UN3373 transport of dangerous goods regulation [40]. Hence, it could facilitate pharmacokinetic sampling as part of drug repurposing trial under stringent risk mitigation strategies. The strong correlation observed between tizoxanide concentration in plasma and DBS, along with good linearity in linear regression analysis allowed reasonable confidence in predicting plasma concentrations from DBS concentrations using the linear equation generated. The strong correlation between plasma concentration and DBS-derived plasma concentration indicated the strong potential of replacing the plasma sampling method of the drug with the DBS sampling approach. Advantages include ease of sample collection, no need for centrifugation, storage at room temperature for stable analytes, and no need for specialized shipment beyond three-component packaging. However, more studies are required to establish room temperature stability, and the impact of other variables not evaluated in the present study on DBS concentration (e.g. hematocrit, and plasma protein binding), in the same way these have been evaluated for some drugs, such as efavirenz and dolutegravir, in the past [41, 42].

## Data availability statement

The data supporting the findings of this study are available upon a request to the corresponding author. No part of the data, apart from the supplementary data, is made publicly available.

