## Supplemental file for "Atazanavir/ritonavir increased tizoxanide exposure from oral nitazoxanide through pharmacokinetic interaction in healthy volunteers"

Supplementary data S1: Accuracy and precision of plasma and DBS analysis

Plasma

| **Nominal conc (ng/mL)** | **Inter-assay** | | | |  | **Intra-assay** | | | |
| --- | --- | --- | --- | --- | --- | --- | --- | --- | --- |
|  | Mean (ng/mL) | SD | %Accuracy | %CV |  | Mean (ng/mL) | SD | %Accuracy | %CV |
| 50 | 46.9 | 3.9 | 84.4 | 8.3 |  | 45.1 | 0.5 | 90.3 | 1.0 |
| 150 | 154.0 | 9.8 | 102.6 | 6.4 |  | 162.1 | 3.5 | 108.0 | 2.2 |
| 400 | 410.7 | 33.8 | 92.4 | 8.2 |  | 448.8 | 16.0 | 112.2 | 3.6 |
| 1000 | 1060.0 | 85.0 | 94.2 | 8.0 |  | 974.6 | 40.9 | 97.5 | 4.2 |
| 2500 | 2671.6 | 196.2 | 106.9 | 7.3 |  | 2640.2 | 165.3 | 105.6 | 6.3 |
| 5000 | 5076.0 | 359.9 | 101.5 | 7.1 |  | 5205.5 | 192.8 | 104.1 | 3.7 |
| 10000 | 9850.4 | 436.9 | 98.5 | 4.4 |  | 9797.1 | 416.5 | 98.0 | 4.3 |
| 15000 | 14055.0 | 1239.5 | 93.7 | 8.8 |  | 14008.4 | 1301.6 | 93.4 | 9.3 |
| 20000 | 18729.4 | 1975.5 | 93.6 | 10.5 |  | 19647.2 | 1158.0 | 98.2 | 5.9 |
| LLOQ (50 ng/mL) | 47.3 | 3.3 | 94.6 | 7.0 |  | 46.6 | 4.5 | 93.1 | 9.7 |
| LQC (120 ng/mL) | 116.9 | 10.6 | 97.4 | 9.0 |  | 125.9 | 9.2 | 104.9 | 7.3 |
| MQC (8600 ng/mL) | 8455.0 | 896.7 | 98.3 | 10.6 |  | 8934.3 | 697.2 | 103.9 | 7.8 |
| HQC (18000 ng/mL) | 18080.8 | 1481.2 | 100.4 | 8.2 |  | 19464.7 | 1058.7 | 108.1 | 5.4 |

Dried blood spot (DBS)

| **Nominal conc (ng/mL)** | **Inter-assay** | | | |  | **Intra-assay** | | | |
| --- | --- | --- | --- | --- | --- | --- | --- | --- | --- |
|  | Mean (ng/mL) | SD | %Accuracy | %CV |  | Mean (ng/mL) | SD | %Accuracy | %CV |
| 50 | 51.6 | 6.25 | 103.1 | 12.12 |  | 54.0 | 4.12 | 107.9 | 4.12 |
| 150 | 145.4 | 14.8 | 97.0 | 10.18 |  | 137.7 | 6.05 | 91.8 | 4.39 |
| 400 | 414.6 | 29.25 | 96.3 | 7.05 |  | 417.9 | 9.22 | 104.5 | 2.21 |
| 1000 | 998.9 | 86.1 | 99.9 | 8.63 |  | 987.83 | 68.29 | 98.8 | 6.91 |
| 2500 | 2418.6 | 236.76 | 96.8 | 9.79 |  | 2487.7 | 93.88 | 99.5 | 3.77 |
| 5000 | 4902.9 | 478.28 | 98.1 | 9.76 |  | 4660.5 | 235.21 | 93.2 | 5.05 |
| 10000 | 9829.3 | 773.53 | 98.3 | 7.87 |  | 9403.2 | 518.96 | 94 | 5.52 |
| 15000 | 15510.0 | 720.3 | 103.4 | 4.64 |  | 14828.3 | 624.43 | 98.9 | 4.21 |
| 20000 | 20058.4 | 673.36 | 100.3 | 3.36 |  | 19944.4 | 97.91 | 99.7 | 0.49 |
| LLOQ (50 ng/mL) | 52.9 | 5.7 | 105.7 | 10.8 |  | 49.5 | 6.5 | 99.1 | 13.1 |
| LQC (120 ng/mL) | 118.3 | 13.5 | 98.6 | 11.4 |  | 117.5 | 13.1 | 97.9 | 11.2 |
| MQC (8600 ng/mL) | 9085.2 | 729.1 | 105.6 | 8.1 |  | 9012.3 | 881.7 | 104.8 | 9.8 |
| HQC (18000 ng/mL) | 17604.7 | 1793.4 | 97.9 | 10.2 |  | 19504.4 | 648.6 | 108.4 | 3.3 |

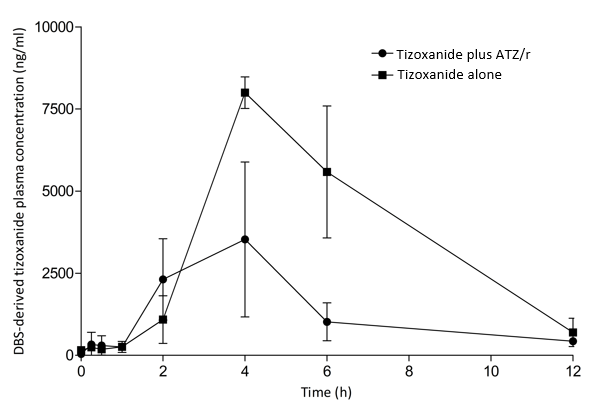

Supplementary S2: DBS-derived plasma concentration-time curve of tizoxanide alone and with atazanavir/ritonavir (ATZ/r); data is presented here as mean (95% CI)

Supplementary S3: Pharmacokinetic parameters of DBS-derived plasma tizoxanide with and without atazanavir/ritonavir

| Pharmacokinetic parameters  (n = 18) | NTZ alone | |  | NTZ plus ATZ/r | | GMR [%, (90% CI)] |
| --- | --- | --- | --- | --- | --- | --- |
|  | Mean (%CV) | GM (%CV) |  | Mean (%CV) | GM (%CV) |  |
| AUC_0-12h_ (ng.h/mL) | 16168.4 (75.8) | 13646.2 (8.9) |  | 42403.0 (24.3) | 41772.6 (2.3) | 3.1 (1.5 – 6.2) |
| C_max_ (ng/mL) | 3528.3 (94.4) | 2626.5 (14.5) |  | 8034.7 (7.8) | 8022.5 (0.9) | 3.05 (1.68 – 10.5) |
| Cτ (ng/m L) | 389.1 (14.2) | 387.1 (2.4) |  | 699.0 (18.3) | 550.5 (5.8) | 1.42 (1.28 – 2.04) |
